# The Political Division toward COVID-19, Vaccines, Contact Tracing Apps, and A Future Pandemic Scenario in the United States: A Survey Result from A National Representative Sample

**DOI:** 10.1101/2023.07.20.23292950

**Authors:** Haijing Hao, Garrett Smith, Yunan Chen, Mainack Mondal, Po-Shen Loh, Staci Smith, Xinru Page

## Abstract

**Objectives:** To investigate the attitudes and behaviors of Americans concerning the COVID-19 pandemic, COVID-19 vaccines, COVID-19 tracing apps, and the actions they believe the government should take during a public health crisis, we designed and conducted a survey during the ongoing COVID-19 emergency.

**Methods:** In January 2022, we administered an online survey on Prolific Academic to 302 participants in the United States, a nationally demographic representative sample. To explore differences in attitudes and opinions among demographic subgroups, we employed several statistical tests, including Mann Whitney U tests, Kruskal-Wallis tests, and chi-squared tests.

**Results:** Our survey results suggest that Americans’ opinions towards the COVID-19 pandemic are severely divided by their political views. There is strong partisan polarization in almost every COVID-19 related question in our survey.

**Policy Implications:** Our findings suggest that policy makers need to consider partisan polarization and the enormous impact it can have on people’s attitudes and behaviors during public health emergencies such as the COVID-19 pandemic. Public health experts need to consider how to convey scientific knowledge about a pandemic without allowing political views to dominate medical conversation.

## 1. Introduction

The COVID-19 pandemic began in early 2020 and has had a huge impact around the world over the past three years.(1,2) The United States is among one of the hardest hit with 102,518,788 confirmed positive cases and 1,110,390 deaths as of Feb. 2, 2023.(3) Public health experts agree that a collective public response is necessary to prevent the spread of this highly contagious virus. (4,5)

However, opinions regarding COVID-19 and COVID-19 vaccines in the United States are divided, largely along partisan lines, or political polarization.(6) Political polarization has been a long standing question in social science research in the U.S. (7) It means that political attitudes sharply divide into two extreme groups, moving away from the center. (8,9) In the United States, Republican-supporting counties have been less likely to maintain social distancing, but are more mobile than Democratic-supporting counties.(10–13) State governors’ recommendations on COVID-19 compliance have been more effective in Democratic-than in Republican-leaning counties.(14) Furthermore, a partisan divide in death rates has been observed, with Republican-majority counties having a higher death rate than those that support the Democratic party.(15,16) Researchers have also uncovered some political polarization toward COVID-19 at the individual level, with liberals engaging more in health protective behaviors than conservatives based on two national surveys in early 2020, at the beginning of the pandemic.(17)

There is also a partisan divide in attitudes towards the COVID-19 vaccine, which is supposed to be another important factor in suppressing the spread of the virus. Vaccines have been a critical tool for human society to combat various infectious diseases in modern times.(18) However, researchers have found that the partisan difference in vaccine hesitancy has increased over time in the U.S.(19) Since the COVID-19 vaccine became available, a study found that counties with a higher percentage of Republican voters had significantly lower vaccination rates, resulting in higher COVID-19 cases and death rates in those counties.(20)

It is important to note that most studies examining partisan differences in the United States, as mentioned above, have used aggregate data such as county-level political vote, county-level COVID-19 death data, or county-level vaccine data, and limited studies of individual-level data on these questions, particularly after the vaccine was available and the pandemic has been a while. Therefore, the present study aims to fill this gap by examining whether an individual’s political views are associated with their opinions, attitudes, and behaviors towards the COVID-19 pandemic and vaccines after two years of the pandemic and one year after the vaccine was available, and whether the results obtained from individual-level data align with the findings of previous aggregate data research. We aim to gain a more comprehensive understanding of how an individual’s political views are linked to their attitudes, opinions, and behaviors concerning the COVID-19 pandemic, COVID-19 vaccines, and COVID-19 tracing apps, and individuals expect what governments should do in a public health crisis. Through this approach, we will be able to provide a more nuanced and robust understanding of the impact of political views on the COVID-19 pandemic at individual level.

## 2. Materials and Methods

### 2.1 Survey design and Data collection

Our survey instrument consisted of three sections that investigated individual opinions and experiences during and about the COVID-19 pandemic, individual’s perceptions about the usefulness of contact tracing in a future pandemic, and their demographics and political views (plus three quality check questions). We deployed our survey on the crowd-sourcing platform Prolific Academic from January 24th, 2022 to January 26th, 2022 to recruit a national representative sample of participants from the U.S. Our study has been approved by IRB category II exempt, and all participants had agreed on the online consent form before they could take the online survey. The national representative here means that a sample reflects the demographic distribution of the U.S. by gender, age, and race to make research findings more generalizable.(21) Earlier studies have shown that participants recruited from Prolific provide high-quality results regarding user perception about software and digital platforms.(22)

### 2.2 Measures

We have categorized these outcomes into four main categories: 1) People’s opinions and behaviors related to COVID-19; 2) People’s opinions regarding the COVID-19 vaccine; 3) People’s personal experiences with COVID-19, COVID-19 vaccines, and tracing apps; 4) People’s expectations of government policies during a public health crisis, or a future pandemic. We have also compared these outcome measures among different demographic subgroups, gender, age, and race, and political view-based subgroups, political views and 2020 presidential vote.

### 2.3 Statistical Methods

Because most of the survey questions were answered by 5-point Likert or 7-point Likert options, the distribution of the ordinal data is non-parametric. Hence, we will conduct non-parametric statistical tests, Mann-Whitney-Wilcoxon tests (also called Wilcoxon rank-sum test), to compare the distribution of two independent samples, and Kruskal-Wallis test to examine the opinion differences among multiple subgroups.(23,24) Wilcoxon rank-sum test and Kruskal-Wallis test does not assume any specific distribution for the data but relies on the rank order of the observations. If the survey answers or opinions are Yes or No, we run Chi-squared tests to examine the differences among different subgroups.(25) All statistical analyses were performed using Stata SE 17.

## 3. Results

### 3.1 Descriptive Statistics

Our survey received a total of 302 responses on Prolific platform, which were nationally represent sample of the United States. Of these, approximately 50% identified as female, 48.34% as male, and 1.66% (5) as other gender. Regarding racial demographics, 8.75% identified as Asian, 15.15% as African American Black, 70.37% as White, and 5.72% as other minorities, such as American Indian, Native Hawaiian, others, or preferred not to say. We categorized respondents into six age groups: 18 to 24 years old (11.1% of respondents), 25 to 34 (20.88%), 35 to 44 (17.51%), 45 to 54 (18.52%), 55 to 64 (19.87%), and 65 years old or above (12.12%). In terms of political views, about 22.19% of respondents identified as Very Liberal, 36.42% as Liberal, 24.50% as Moderate, 11.26% as Conservative, 3.97% as Very Conservative, and 1.66% preferred not to say. We also categorized the respondents by their presidential vote in 2020. In our sample, 12.91% voted for Trump in 2020 presidential election, 67.88% voted for Biden, 11.92% not vote, and 7.28% voted for others. Based on our survey results, approximately 81% of respondents reported receiving a COVID-19 vaccine, and around 18% reported having tested positive for COVID-19, as of Jan. 2022, two years after the pandemic.

### 3.2 Statistical Analysis

For the present study, we conducted statistical analysis among different subgroups by gender, age, race, political views, and the 2020 presidential vote. The respondents who answered other gender or chose not to disclose their gender, age, or political views were dropped from our statistical analysis because the number is too small to have statistical significance. Regarding the 2020 presidential vote, the present study focuses on the two groups, which voted for Biden and voted for Trump.

Table 1 presents the statistical analysis of nine questions regarding people’s opinions and behaviors concerning the COVID-19 pandemic. The responses to all questions were measured on a five-point Likert scale, ranging from “Disagree Strongly” to “Agree Strongly,” with values from 1 to 5. The results reveal that the median scores for all questions were either 4 or 5, indicating that approximately half of the respondents somewhat agreed or strongly agreed with the statements. Regarding social compliance behaviors, the majority of respondents reported practicing mask-wearing, social distancing, and avoiding gatherings. Concerning COVID-19, respondents expressed apprehension about contracting the virus, hospital capacities, the impact on the economy, the virus’s dangerousness, and the potential for new variants. Lastly, respondents believed that governments should take additional measures to prevent the spread of COVID-19.

**Table 1.** Statistical analysis of Americans’ social compliance behaviors, concerns about COVID-19, and government roles.

|  | When I leave the house, I always maintain social distance.<br>(Med./Mean) | I wear my mask whenever I leave the house because of COVID-19.<br>(Med./Mean) | I avoid events or social gatherings with other people because of COVID-19.<br>(Med./Mean) | I am very worried about getting COVID-19.<br>(Med./Mean) | I am concerned about the hospital capacity during the COVID-19 pandemic.<br>(Med./Mean) | I believe that COVID-19 is more dangerous than the common flu.<br>(Med./Mean) | I worry about the effects of variants of the disease.<br>(Med./Mean) | I am very worried about the effect of the pandemic on the economy.<br>(Med./Mean) | I believe that my government should do more to deal with COVID-19.<br>(Med./Mean) |
| --- | --- | --- | --- | --- | --- | --- | --- | --- | --- |
| **All** | 4/3.97 | 4/4.13 | 4/4 | 4/3.35 | 5/4.30 | 5/4.43 | 5/4.27 | 4/4.05 | 4/3.93 |
| Female | 4/4.05 | 5/4.33 | 4/4.06 | 4/3.56 | 5/4.46 | 5/4.56 | 5/4.42 | 4/4.11 | 4/4.11 |
| Male | 4/3.86 | 4/3.88 | 4/3.90 | 3/3.12 | 5/4.12 | 5/4.30 | 4/4.12 | 4/4.02 | 4/3.93 |
| **Wilcoxon rank-sum test** | 0.2134 | 0.0009*** | 0.6704 | 0.0062** | 0.0035** | 0.0083** | 0.0157* | 0.4988 | 0.0039** |
| Age 18-24 | 4/3.68 | 5/4.12 | 4/3.71 | 4/3.35 | 5/4.41 | 4/4.18 | 5/4.26 | 4/3.85 | 4/3.88 |
| Age 25-34 | 4/3.75 | 4/4.03 | 4/3.70 | 4/3.09 | 4.5/4.20 | 5/4.19 | 4/4.11 | 4/3.97 | 4/3.95 |
| Age 35-44 | 4/4.07 | 5/4.37 | 5/4.26 | 4/3.63 | 5/4.65 | 5/4.69 | 5/4.50 | 4/4.00 | 5/4.28 |
| Age 45-54 | 4/3.95 | 4/3.84 | 4/3.87 | 3/3.29 | 5/4.11 | 5/4.44 | 4/4.20 | 4/4.15 | 4/3.91 |
| Age 55-64 | 5/4.25 | 5/4.25 | 5/4.25 | 4/3.56 | 5/4.29 | 5/4.64 | 5/4.27 | 4/4.12 | 4/3.73 |
| Age 65+ | 4/4.06 | 5/4.22 | 5/4.14 | 4/3.14 | 5/4.14 | 5/4.36 | 5/4.36 | 5/4.19 | 4/3.78 |
| **Kruskal-Wallis test** | 0.0282* | 0.1654 | 0.0002** | 0.1924 | 0.0952 | 0.0101* | 0.4623 | 0.5397 | 0.2360 |
| Asian | 4/3.62 | 4.5/4.23 | 4/3.69 | 3/3.04 | 4/3.92 | 4.5/4.31 | 4/4.08 | 4/4.08 | 3/3.35 |
| African American | 5/4.29 | 5/4.49 | 5/4.20 | 4/3.56 | 5/4.38 | 5/4.51 | 5/4.44 | 5/4.26 | 4/3.96 |
| White | 4/3.94 | 5/4.02 | 4/3.99 | 4/3.35 | 5/4.33 | 5/4.45 | 5/4.29 | 4/4.01 | 4/4.03 |
| Others | 4/4.05 | 5/4.32 | 4/4.00 | 4/3.26 | 5/4.32 | 5/4.16 | 4/3.95 | 4/3.90 | 4/3.58 |
| **Kruskal-Wallis test** | 0.0097** | 0.0226* | 0.2446 | 0.4115 | 0.1229 | 0.3161 | 0.2215 | 0.4974 | 0.0220* |
| Very Liberal | 4/4.28 | 5/4.49 | 5/4.36 | 4/3.67 | 5/4.67 | 5/4.85 | 5/4.60 | 4/3.67 | 5/4.63 |
| Liberal | 4/4.27 | 5/4.44 | 4/4.31 | 4/3.75 | 5/4.58 | 5/4.76 | 5/4.60 | 4/3.97 | 4/4.22 |
| Moderate | 4/3.76 | 4/3.93 | 4/3.64 | 3/3.04 | 4/4.16 | 4/4.05 | 4/3.96 | 5/4.37 | 4/3.47 |
| Conservative | 3/3.21 | 3.5/3.21 | 4/3.21 | 2/2.32 | 3.5/3.35 | 4/3.74 | 4/3.59 | 5/4.24 | 3/3.12 |
| Very Conservative | 2.5/2.92 | 2/2.67 | 3/3.17 | 2/2.42 | 3/2.92 | 4/3.33 | 3.5/3.17 | 4.5/4.50 | 2.5/2.58 |
| **Kruskal-Wallis test** | 0.0001*** | 0.0001*** | 0.0001*** | 0.0001*** | 0.0001*** | 0.0001*** | 0.0001*** | 0.0095** | 0.0001*** |
| Vote for Biden in 2020 | 4/4.19 | 5/4.40 | 4/4.24 | 4/3.64 | 5/4.64 | 5/4.73 | 5/4.55 | 4/3.91 | 5/4.25 |
| Vote for Trump in 2020 | 3/3.05 | 3/3.00 | 4/3.05 | 2/2.10 | 3/2.82 | 4/3.41 | 4/3.23 | 5/4.54 | 3/2.85 |
| **Wilcoxon rank-sum test** | <0.0001*** | <0.0001*** | <0.0001*** | <0.0001*** | <0.0001*** | <0.0001*** | <0.0001*** | 0.0008*** | <0.0001*** |

Note: \* denotes statistical significance at the 5% level. \*\* denotes statistical significance at the 1% level. \*\*\* denotes statistical significance at the 0.1% level.

Due to space limitations, Table 1 only displays the statistical analysis comparing the median and mean values of different demographic and political subgroups. However, if needed, we can provide the complete distributions of each subgroup. Overall, significant differences were observed in some responses based on gender, age, race, political views, and the 2020 presidential votes.

Six out of the nine questions exhibited statistically different opinions between women and men. Females indicated higher agreement in wearing masks when outside the house (median 5 vs. median 4, mean 4.33 vs. mean 3.88, Wilcoxon test p-value = 0.0009), greater concern about contracting COVID-19 (median 4 vs. median 3, mean 3.56 vs. mean 3.12, Wilcoxon test p-value = 0.0062), heightened worry about hospital capacities during the pandemic (mean 4.46 vs. mean 4.12, Wilcoxon test p-value = 0.0035), stronger belief that COVID-19 is more dangerous than the common flu (mean 4.56 vs. mean 4.30, Wilcoxon test p-value = 0.0083), increased concern about the effects of variants (median 5 vs. median 4, mean 4.42 vs. mean 4.12, Wilcoxon test p-value = 0.0157), and a greater belief that the government should take further action to address COVID-19 (mean 4.11 vs. mean 3.93, Wilcoxon test p-value = 0.0039). Although these six questions exhibited statistically different average opinion scores, the magnitude of the differences was not substantial, ranging from 3.12 to 4.56, all leaning toward agreement.

Different age groups demonstrated diverse opinions on three of the nine questions: maintaining social distance (Kruskal-Wallis test p-value = 0.0282), avoiding social gatherings (Kruskal-Wallis test p-value = 0.0002), and perceiving COVID-19 as more dangerous than the common flu (Kruskal-Wallis test p-value = 0.0101). Among various racial subgroups, differing opinions were evident on three questions: maintaining social distance (Kruskal-Wallis test p-value = 0.0097), wearing masks when outside the house (Kruskal-Wallis test p-value = 0.0226), and the belief that governments should take further action (Kruskal-Wallis test p-value = 0.0220). However, overall, these differences were not substantial, as the opinions qualitatively aligned among different age or racial subgroups.

Nevertheless, significant differences were observed in all nine questions when considering political view-based subgroups or 2020 presidential vote subgroups. Quantitatively, the opinion scores exhibited a linear correlation with political views. The more liberal the subgroup, the greater their concerns about COVID-19, and vice versa. Furthermore, significant disparities were observed on four questions, with the Very Liberal group exhibiting a median value of 2 compared to the Very Conservative group’s median value of 4 or 5. Similar contrasting differences were observed in average scores between the two groups, with mean scores lower than 3 leaning toward disagreement for one group, and mean scores close to 5 leaning toward strong agreement for the other. The Very Liberal subgroup expressed stronger agreement than the Very Conservative subgroup regarding maintaining social distance when outside the house (median 4 vs. median 2.5, mean 4.28 vs. mean 2.92, Kruskal-Wallis test p-value = 0.0001), wearing masks when outside the house (median 5 vs. median 2, mean 4.49 vs. mean 2.67, Kruskal-Wallis test p-value = 0.0001), concern about contracting COVID-19 (median 4 vs. median 2, mean 3.67 vs. mean 2.42, Kruskal-Wallis test p-value = 0.0001), and the belief that governments should take further action to address COVID-19 (median 5 vs. median 2.5, mean 4.63 vs. mean 2.258, Kruskal-Wallis test p-value = 0.0001). The differences between the Vote for Biden and Vote for Trump groups followed similar patterns as the political view-based subgroups, showcasing a partisan division on all nine questions, with Biden supporters expressing stronger agreement regarding COVID-19 issues, while Trump supporters exhibited stronger disagreement.

Table 2 presents three questions regarding COVID-19 vaccines, with responses provided on a five-point Likert scale. The median and mean scores for all respondents range from 3.5 to 5, indicating that approximately half of the respondents agreed, on average, with the three vaccine-related questions. Among the gender subgroups, only one question displayed a statistically significant difference in opinion, specifically regarding whether individuals who have received the vaccine should still exercise caution. Women showed stronger agreement than men on this question (mean 4.74 vs. mean 4.49, Wilcoxon test p-value = 0.0009). There were no statistically significant differences among the various age subgroups for the three vaccine-related questions. Among the four racial subgroups, the only question with statistically different opinions was whether vaccinated individuals should remain cautious (Kruskal-Wallis test p-value = 0.0056). However, the median or mean scores for each subgroup fell between 4.50 and 5, all indicating strong agreement.

**Table 2.** Americans’ opinions about the COVID-19 vaccine.

|  | I believe that getting the vaccine should be mandatory. (Median/Mean) | I believe that the vaccine works. (Median/Mean) | I think that those who have received the vaccine should still be careful. (Median/Mean) |
| --- | --- | --- | --- |
| **All** | 4/3.5 | 5/4.62 | 5/4.27 |
| Female | 4/3.69 | 5/4.27 | 5/4.74 |
| Male | 4/3.28 | 5/4.26 | 5/4.49 |
| **Wilcoxon test** | 0.0273* | 0.6650 | 0.0009*** |
| Age 18-24 | 4/3.47 | 5/4.26 | 5/4.32 |
| Age 25-34 | 3.5/3.36 | 4.5/4.11 | 5/4.64 |
| Age 35-44 | 4/3.83 | 5/4.52 | 5/4.70 |
| Age 45-54 | 3/3.09 | 5/3.96 | 5/4.55 |
| Age 55-64 | 4/3.73 | 5/4.44 | 5/4.68 |
| Age 65+ | 4/3.53 | 5/4.39 | 5/4.75 |
| **Kruskal–Wallis test** | 0.2303 | 0.0896 | 0.2184 |
| Asian | 4/3.77 | 5/4.31 | 5/4.50 |
| African American | 4/3.38 | 4/3.96 | 5/4.87 |
| White | 4/3.53 | 5/4.36 | 5/4.56 |
| Others | 3/3.11 | 5/4.00 | 5/4.84 |
| **Kruskal–Wallis test** | 0.6571 | 0.1685 | 0.0056** |
| Very Liberal | 5/4.25 | 5/4.78 | 5/4.79 |
| Liberal | 4/4.04 | 5/4.63 | 5/4.74 |
| Moderate | 3/2.88 | 4/3.85 | 5/4.57 |
| Conservative | 2/2.15 | 4/3.35 | 4/4.03 |
| Very Conservative | 2/2.17 | 4/3.33 | 5/4.50 |
| **Kruskal–Wallis test** | 0.0001*** | 0.0001*** | 0.0001*** |
| Vote for Biden | 5/4.07 | 5/4.67 | 5/4.72 |
| Vote for Trump | 1/1.62 | 3/3.10 | 4/4.21 |
| **Wilcoxon test** | <0.0001*** | <0.0001*** | 0.0002*** |

Once again, the political view-based subgroups exhibited significantly different opinions on all three vaccine questions, with all Kruskal-Wallis test p-values equal to 0.0001. The more liberal the subgroup, the stronger their belief in the vaccine’s effectiveness (Very Liberal subgroup’s median = 5 and mean = 4.78 vs. Very Conservative subgroup’s median = 4 and mean = 3.33, Kruskal-Wallis test p-value = 0.0001), and their support for mandatory vaccination (Very Liberal subgroup’s median = 5 and mean = 4.25 vs. Very Conservative subgroup’s median = 2 and mean = 2.17, Kruskal-Wallis test p-value = 0.0001), and their support for still being cautious after vaccination (Very Liberal subgroup’s mean = 4.79 vs. Very Conservative subgroup’s mean = 4.50, Kruskal-Wallis test p-value = 0.0001). The opinion scores exhibited a wide range, spanning from 2 on the disagreement side to 5 on the strong agreement side, representing two opposite ends. A similar pattern was observed among the two 2020 presidential vote groups, with both groups expressing more extreme opinions compared to the Very Liberal and Very Conservative groups on all three questions.

Table 3 presents four outcomes related to individuals’ personal experiences with COVID-19, vaccination, accessibility to the vaccine, and usage of a tracing app. The responses to these four questions were binary: Yes or No. It was found that 98% of people reported having access to the vaccine, and 81% reported having been vaccinated. Approximately 18% of individuals answered Yes when asked if they had tested positive for COVID-19. This percentage aligns with the national test positivity rate observed around January 2022 (26). Furthermore, approximately 15% of respondents reported having used a tracing app.

**Table 3.** Americans’ experiences with COVID-19, COVID-19 vaccine, and COVID-19 tracing apps.

|  | I have tested positive for COVID-19 in the past.<br>(Median/Mean) | I have been vaccinated.<br>(Median/Mean) | Do you have access to the COVID-19 vaccine?<br>(Median/Mean) | During pandemics such as with COVID-19, public health departments have used contact tracing apps to slow the spread of the disease. The mobile application usually does this by identifying individuals that may have been in contact with an infected person. Have you ever used a contact tracing app? (Median/Mean) |
| --- | --- | --- | --- | --- |
| **All** | 0/0.18 | 1/0.81 | 1/0.98 | 0/0.15 |
| Female | 0/0.19 | 1/0.81 | 1/0.97 | 0/0.12 |
| Male | 0/0.16 | 1/0.83 | 1/0.99 | 0/0.19 |
| **Chi-squared test** | 0.434 | 0.642 | 0.188 | 0.131 |
| Age 18-24 | 0/0.29 | 1/0.88 | 1/0.97 | 0/0.29 |
| Age 25-34 | 0/0.23 | 1/0.78 | 1/0.98 | 0/0.15 |
| Age 35-44 | 0/0.28 | 1/0.85 | 1/0.98 | 0/0.17 |
| Age 45-54 | 0/0.09 | 1/0.69 | 1/0.98 | 0/0.11 |
| Age 55-64 | 0/0.07 | 1/0.85 | 1/0.98 | 0/0.12 |
| Age 65+ | 0/0.11 | 1/0.89 | 1/0.97 | 0/0.11 |
| **Chi-squared test** | 0.004** | 0.097 | 0.996 | 0.232 |
| Asian | 0/0.08 | 1/0.92 | 1/1 | 0/0.27 |
| African American | 0/0.11 | 1/0.82 | 1/1 | 0/0.12 |
| White | 0/0.2 | 1/0.81 | 1/0.98 | 0/0.16 |
| Others | 0/0.21 | 1/0.68 | 1/0.95 | 0/0 |
| **Chi-squared test** | 0.267 | 0.241 | 0.451 | 0.091 |
| Very Liberal | 0/0.18 | 1/0.93 | 1/0.99 | 0/0.30 |
| Liberal | 0/0.17 | 1/0.90 | 1/1.00 | 0/0.19 |
| Moderate | 0/0.16 | 1/0.7 | 1/0.97 | 0/0.06 |
| Conservative | 0/0.12 | 1/0.62 | 1/0.94 | 0/0 |
| Very Conservative | 0/0.42 | 1/0.67 | 1/1.00 | 0/0 |
| **Chi-squared test** | 0.221 | <0.0001*** | 0.185 | <0.0001*** |
| Vote for Biden | 0/0.17 | 1/0.94 | 1/1.00 | 0/0.20 |
| Vote for Trump | 0/0.21 | 1/0.51 | 1/0.97 | 0/0 |
| **Chi-squared test** | 0.551 | <0.0001*** | 0.187 | 0.003** |

The experience of testing positive for COVID-19 exhibited a statistically significant difference across age subgroups (Chi-squared test p-value = 0.004), ranging from 7% in the 55 to 64 years old group to 29% in the 18 to 24 years old group. However, no significant differences were observed for any other subgroups. Political view-based subgroups displayed statistically different responses to the question of vaccination (Chi-squared test p-value < 0.0001). The Very Liberal subgroup reported a vaccination rate of 93%, while the Conservative and Very Conservative subgroups reported vaccination rates of 62% to 67%. Similarly, a higher likelihood of using a tracing app was observed among more liberal subgroups (Very Liberal group’s mean = 0.3 and Liberal group’s mean = 0.19 vs. Very Conservative group’s mean = 0 and Conservative group’s mean = 0, with Chi-squared test p-value < 0.0001). A similar pattern was found among the 2020 presidential vote groups, with Biden supporters more likely to have been vaccinated compared to Trump supporters (Vote for Biden mean = 0.94 vs. Vote for Trump mean = 0.51, Chi-squared test p-value < 0.0001), and Biden supporters being more likely to have used a contact tracing app compared to Trump supporters (Vote for Biden mean = 0.2 vs. Vote for Trump mean = 0, Chi-squared test p-value = 0.003). However, it is worth noting that even the highest rate of tracing app usage by the Very Liberal subgroup, was only 30%, which is relatively low. Additionally, none of the individuals in the two conservative subgroups reported having ever used a tracing app.

It is important to highlight that the answers to the two fact-based questions, “I have tested positive in the past” and “Do you have access to the COVID-19 vaccine,” showed no statistically significant differences among any groups. This indicates that testing positive or having access to the vaccine is not related to any specific demographics or political views. The vaccine was nearly equally accessible to all subgroups, ranging from 94% to 100%.

Table 4 presents the responses to five survey questions regarding the public’s perspectives on what type of information should be shared by local governments to instill a sense of health security in the event of a future pandemic. These questions form part of a hypothetical scenario designed to gauge the common expectations during a potential public health crisis. They include questions about the known count or percentage of diagnosed individuals, the progression trend of new cases, the vaccination rate, and the frequency of self-diagnosis of the infection. Responses were captured using a seven-point Likert scale that ranged from ‘Disagree Completely’ (1) to ‘Agree Completely’ (7). For all five questions, the median score was 6, while the mean scores hovered near 6, implying that approximately half of the respondents, or on average, agreed substantially that the dissemination of certain disease-related data by their local government would contribute to their feeling of health security.

**Table 4.** Americans’ opinions on what information governments should share if a future pandemic occurs.

|  | Knowing how many people have been diagnosed with the disease in my county, city, and local school in the last 24 hours would enable me to protect my health.<br>(Median/Mean) | Knowing what proportion of people have been diagnosed with the disease in my county, city, and local school in the last 24 hours would enable me to protect my health.<br>(Median/Mean) | Knowing whether the number of new cases in my county is trending upwards or downwards, and by how much, would enable me to protect my health.<br>(Median/Mean) | Knowing what percentage of people have been vaccinated against the disease in my state would enable me to protect my health.<br>(Median/Mean) | Knowing when others have self-diagnosed that they have this disease (with 80% certainty) would enable me to protect my health.<br>(Median/Mean) |
| --- | --- | --- | --- | --- | --- |
| **All** | 6/5.57 | 6/5.59 | 6/5.6 | 6/5.23 | 6/5.92 |
| Female | 6/5.74 | 6/5.77 | 6/5.85 | 6/5.41 | 6/6.05 |
| Male | 5/5.39 | 5/5.38 | 5/5.30 | 5/5.01 | 6/5.78 |
| **Wilcoxon test** | 0.0134* | 0.0163* | 0.0004*** | 0.0416* | 0.0141* |
| Age 18-24 | 6.5/5.85 | 7/5.91 | 7/5.97 | 6/5.50 | 7/5.82 |
| Age 25-34 | 6/5.78 | 6/5.72 | 6/5.72 | 6/5.23 | 6/6.6 |
| Age 35-44 | 6/5.74 | 6/5.78 | 6/5.57 | 6/5.63 | 6/5.76 |
| Age 45-54 | 5/5.20 | 5/5.18 | 5/5.27 | 5/4.85 | 6/5.62 |
| Age 55-64 | 5/5.36 | 6/5.51 | 6/5.51 | 5/4.97 | 6/6.05 |
| Age 65+ | 6/5.58 | 6/5.56 | 6/5.69 | 6/5.33 | 7/6.11 |
| **Kruskal–Wallis test** | 0.0249* | 0.0311* | 0.0420* | 0.1089 | 0.0830 |
| Asian | 5/5.12 | 5/5.31 | 5.5/5.42 | 6/5.31 | 6/5.81 |
| African American | 6/5.78 | 6/5.82 | 6/5.87 | 5/5.33 | 6/5.98 |
| White | 6/5.53 | 6/5.53 | 6/5.5 | 5/5.12 | 6/5.93 |
| Others | 7/6.11 | 6/6.11 | 7/6.21 | 6-Jul | 7/5.90 |
| **Kruskal–Wallis test** | 0.0277* | 0.0975 | 0.0809 | 0.1579 | 0.5913 |
| Very Liberal | 6/5.97 | 6/5.99 | 6/5.87 | 6/5.69 | 7/6.36 |
| Liberal | 6/5.82 | 6/5.76 | 6/5.81 | 6/5.47 | 6/6.12 |
| Moderate | 6/5.53 | 6/5.61 | 6/5.65 | 5.5/5.26 | 6/5.81 |
| Conservative | 4/4.24 | 4.5/4.41 | 5/4.41 | 4/4.06 | 5/4.94 |
| Very Conservative | 5/4.92 | 5/5.00 | 5.5/5.00 | 3.5/3.33 | 5.5/5.33 |
| **Kruskal–Wallis test** | 0.0001*** | 0.0001*** | 0.0001*** | 0.0001*** | 0.0001*** |
| Vote for Biden | 6/5.81 | 6/5.80 | 6/5.80 | 6/5.57 | 7/6.15 |
| Vote for Trump | 4/4.28 | 4/4.41 | 5/4.54 | 4/3.54 | 5/5.33 |
| **Wilcoxon test** | <0.00001*** | <0.00001*** | <0.00001*** | <0.00001*** | 0.0001*** |

An interesting gender difference emerged in the responses to all five questions. Females expressed stronger agreement than males about the impact of such information on their health security. This difference was statistically significant, as confirmed by the Wilcoxon test, for all the following topics: the number of local diagnoses (female median = 6, mean = 5.74 vs. male median = 5, mean = 5.39, with Wilcoxon test p-value = 0.0134), the proportion of local diagnoses (female median = 6, mean = 5.77 vs. male median = 5, mean = 5.38, with Wilcoxon test p-value = 0.0163), the trend of new cases (female median = 6, mean = 5.85 vs. male median = 5, mean = 5.30, with Wilcoxon test p-value = 0.0004), the local vaccination rate (female median = 6, mean = 5.41 vs. male median = 5, mean = 5.01, with Wilcoxon test p-value = 0.0416), and the prevalence of self-diagnosis (female median = 6, mean = 6.05 vs. male median = 6, mean = 5.78, with Wilcoxon test p-value = 0.0141).

Statistically significant differences also arose among different age groups for three of the five questions: the number of local diagnoses (Kruskal–Wallis test = 0.0249), the proportion of local diagnoses (Kruskal–Wallis test = 0.0311), and the trend of new cases (Kruskal–Wallis test = 0.0420). In terms of racial differences, only one question yielded a significant difference of opinion - the number of people diagnosed locally (Kruskal–Wallis test = 0.0277).Again, political orientation played a decisive role in shaping the responses to all five questions (all Kruskal-Wallis test p-values were 0.0001). The question regarding the public’s perception of how a high vaccination rate could protect their health proved to be particularly polarizing.

Respondents identifying as ‘Very Liberal’ had a median score of 6 and mean score of 5.69, while those identifying as ‘Very Conservative’ scored a median of 3.5 and a mean of 3.33, reflecting differing beliefs about the role of vaccination in pandemic management. Similarly, patterns emerged among 2020 presidential voting groups, with the ‘Vote for Biden’ group demonstrating greater agreement with all proposed protective measures to be shared by the government than the ‘Vote for Trump’ group, and all Kruskal-Wallis tests are statistically significant with p values less than 0.0001.

## 4. Discussion

Studies have underscored the existence of partisan polarization in response to the COVID-19 pandemic in the U.S. A Pew survey spanning thirteen countries identified that American attitudes towards COVID-19 were considerably more politicized than those in other advanced economies. (6) Within the U.S., counties favoring the Republican candidate, Donald Trump, over the Democratic candidate, Hillary Clinton, in the 2016 presidential election, manifested 14% less physical distancing at the onset of the COVID-19 outbreak, in early 2020. (10) Furthermore, another study in 2021 showed that even when holding other factors constant, Republican supporters displayed a 27.8% greater likelihood of mobility than Democratic supporters.(12) These trends are reflected in our survey data, where significant partisan differences at an individual level are observable. For instance, as indicated in Table 1, individuals within the Very Conservative subgroup expressed stronger disagreement than those in the Very Liberal subgroup, on whether they consistently maintain social distancing when venturing outside their homes (median score = 2.5 and mean score = 2.92 vs. median score = 5 and mean score = 4.28, Kruskal-Wallis test p-value =0.0001) and on whether they avoid social gatherings with other people (median score = 3 and mean score = 3.17 vs. median score = 5 and mean score = 4.36, Kruskal-Wallis test p-value =0.0001). One study indicated that, during the initial phase of the COVID-19 outbreak—a critical period for the virus’s proliferation—states governed by Republican leaders were slower to enforce social distancing policies compared to those led by Democratic leaders, even after accounting for other factors.(13) Our data corroborate this finding, showing that, on average, individuals in the most conservative subgroup disagreed with the notion that the government should intensify efforts to manage the COVID-19 pandemic, while their counterparts in the most liberal subgroup advocated for more robust government action (median score = 2.5 and mean score = 2.58 vs. median score 5 and mean score = 4.63, Kruskal-Wallis test p-value =0.0001). In anticipation of a future pandemic, our survey respondents indicated a desire for their local governments to keep them updated on data such as the number/proportion of people diagnosed with the disease, new case trends, the proportion of vaccinated individuals, and the ability of people to self-diagnose in their country, as Table 4 showed. In general, all groups agreed that their local governments should share more data which would protect their own health with median and mean scores above 5 or 6, Agree Mostly or Agree Completely. However, those in the Very Conservative subgroup demonstrated lower support for these data transparency policies compared to the Very Liberal subgroup. The greatest divergence in views concerned the utility of protective measures such as knowing the percentage of people vaccinated (median score = 3.5 and mean score = 3.33 vs. median score = 6 and mean score =5.69, Kruskal-Wallis test p-value = 0.0001). This discrepancy aligns with the observed partisan variance towards COVID-19 vaccines, a subject we will delve into further.

Vaccine hesitancy, a politically polarizing issue in the U.S., has amplified during the COVID-19 pandemic. Historically, effective vaccines have halted the spread of most infectious diseases in modern human history. However, even prior to the COVID-19 pandemic, vaccine hesitancy was a politically charged issue in the U.S., with some Republican political figures publicly opposing vaccines. (27) This partisan split in opinions towards COVID-19 vaccines is even starker. In counties with higher proportions of Republican voters, vaccination rates for COVID-19 were markedly lower, correlating with a higher COVID-19 positive rate and mortality rate. (20) Over time, this partisan polarization in vaccine hesitancy has grown. (19) As Table 2 showed, our survey data spotlight a significant disparity between the liberal subgroups, who generally believe in the vaccine’s efficacy and support mandatory vaccination, and the conservative subgroups, who exhibited greater skepticism towards vaccine effectiveness (median score = 5 and mean score = 4.78 vs median score = 4 and mean score = 3.33, with Kruskal-Wallis test p-value = 0.0001) and were less likely to endorse mandatory vaccination policies (median score = 5 and mean score = 4.25 vs. median score = 2 and mean score = 2.17, with Kruskal-Wallis test p-value = 0.0001).

Digital contact tracing is another crucial tool for controlling the COVID-19 pandemic, even in the wake of available vaccines. (28) Research suggests that digital tracing has proven to be a successful strategy in managing Ebola and tuberculosis epidemics. (29–31) Simulation studies also demonstrate the potential for COVID-19 contact tracing apps to manage the pandemic, contingent on high adoption rates. (32) However, the deployment of digital contact tracing apps has encountered challenges, including limited smartphone penetration, varying acceptance rates for tracing apps, and privacy concerns, among others. (33–35) A study in 2020 identified significant partisan discrepancies in attitudes towards digital contact tracing, with more Republicans expressing opposition (39%) than Democrats (27%) in the early stages of the pandemic. (28) Our 2022 survey data, collected two years into the pandemic at a time when COVID-19 vaccines and contact tracing apps were most accessible, uncovers persistent partisan divides at the individual level. Notably, as Table 3 showed, none of the Conservative or Very Conservative subgroups had ever used a COVID-19 contact tracing app, whereas approximately 30% of Very Liberal subgroup members and about 19% of Liberal subgroup members had utilized such an app. Even though liberal subgroups show a higher adoption rate for contact tracing apps than conservative subgroups, the overall adoption rate remains insufficient for the apps to function effectively. This should alert public health experts to the persistently low acceptance rates of tracing apps in the U.S.

Additionally, the survey data presented in Table 3 reveals that for fact-based questions—such as testing positive for COVID-19 or accessibility to the COVID-19 vaccine—there is no significant difference among individuals with varying political views, which suggests that the reality of the U.S. as well as a good quality of our survey data.

## 5. Conclusions and Limitations

Our study offers several contributions. First, we explore the existence of partisan disparities in attitudes towards the COVID-19 pandemic in the U.S. at an individual level. This investigation utilizes a nationally representative sample obtained in 2022, during the height of the pandemic, from the reputable research-focused online platform, Prolific. The findings derived from our dataset align with previous studies that relied on aggregated county-level data; however, our dataset offers the added advantage of being individual-based, making it more practical from a public policy standpoint. This data affords public health professionals, government officials, and policymakers an alternative perspective to understand the divergence in opinions that is rooted in political views. Second, our survey extends beyond just capturing people’s experiences and views on the COVID-19 pandemic; it also includes hypothetical questions concerning a future pandemic. Last, our research is among the first to scrutinize partisan polarization during the COVID-19 pandemic at the individual level, assessing opinions on COVID-19, vaccine hesitancy, and trust in COVID-19 tracing apps in the U.S.

Despite its contributions, this study is not without limitations. First, our survey’s sample size is relatively small, consisting of approximately 302 participants. Although Prolific has ensured national representation by incorporating a proper proportionate mix of genders, ages, and races, the sample might still fall short of adequately representing a vast and diverse country like the U.S. Consequently, researchers are advised to interpret the data and results with caution. Second, our survey questions might not encompass all relevant facets of the COVID-19 pandemic, reflecting only specific perspectives of Americans concerning the pandemic. Last, as is the case with many survey studies, respondents might have exhibited reluctance in divulging their views on sensitive questions, despite assurances of anonymity.

## 6. Public Health Implications

Our survey data underscores a significant partisan polarization in the U.S. concerning COVID-19, its vaccines, and tracing apps. Political divisions surface in nearly every COVID-19-related question within our survey. Consequently, we suggest four policy implications. First, public health administrators and researchers should contemplate strategies to reduce partisan differences concerning the COVID-19 pandemic or any future pandemics and epidemics. Emphasizing science-based guidance from medical experts is crucial to ensure appropriate pandemic responses. Second, political figures may need to moderate their influence within the realm of public health. This restraint can help prevent health decisions driven more by partisan bias than by public well-being. Third, it falls upon public health experts and technologists to enhance their communication about the effectiveness and security of digital tracing apps. Clear messaging about these apps’ capacity to control infectious diseases is necessary. Last, people’s belief that data availability and transparency aids their health protection during a pandemic necessitates consideration. Therefore, governments should deliberate on making data reflecting local conditions easily accessible.

## Data Availability

Please contact Prof. Xinru Page for the data availability.

